# Assessing healthcare professionals’ experiences of delivering opportunistic, weight-related conversations in a mental health setting: a mixed methods study

**DOI:** 10.64898/2025.12.16.25342378

**Authors:** Angela M. Rodrigues, Emma Kemp, Sally Faulkner, Kate McBride, Maria Raisa Jessica Aquino, Rob Wilson, Milica Vasiljevic, Craig Robson, Jill Harland, Mish Loraine, Catherine Haighton

**Affiliations:** Northumbria University, Newcastle upon Tyne, NE7 7XA, UK; University of Sheffield, Sheffield, S10 2TN, UK; Cumbria, Northumberland, Tyne & Wear NHS Foundation Trust, St Nicholas Hospital, Newcastle upon Tyne, NE3 3XT, UK; Newcastle University, Newcastle Upon Tyne, NE1 4AX, UK; Manchester Metropolitan University, Manchester M15 6BX, UK; Durham University, Durham, DH1 3LE, UK; Northumbria Healthcare NHS Foundation Trust, North Tyneside General Hospital, NE29 8NH, UK

**Keywords:** public mental health, training implementation, behaviour change, health services research

## Abstract

**Background:** Making Every Contact Count (MECC) is a person-centred approach to health behaviour change, utilising behavioural science to promote healthy lifestyle choices. Weight-related conversations in mental health settings are particularly important, with users noting gaps in information on medication-related weight gain and support needs. MECC training can address these gaps by improving staff confidence and service delivery, enhancing weight management for individuals with serious mental illness. This study explores staff experiences of weight-related MECC conversations in a mental health setting.

**Methods:** This mixed-methods study involved healthcare staff from a National Health Service (NHS) mental health inpatient setting in Northeast England. A quantitative online survey administered pre- MECC training, post-MECC training, and 8-10 weeks later captured staff perspectives. Qualitative interviews were conducted with trained and non-trained staff to explore MECC implementation. Quantitative data were analysed using independent and paired-samples t-tests, while qualitative data underwent thematic analysis, using the COM-B framework that examines Capability, Opportunity, and Motivation as key drivers of behaviour.

**Results:** Thirty-six staff completed the pre-training survey, 20 completed the post-training survey, and 25 participated in interviews (15 trained, 10 non-trained staff). Quantitative analysis showed that training improved staff perceptions of confidence and motivation to deliver MECC, though most changes were not statistically significant. Perceptions around opportunity (time, resources, social support) declined at follow-up. Perceived importance and usefulness of MECC also declined over time (p < .05). Qualitative analysis identified barriers and facilitators, mapped to the COM-B model. Trained staff highlighted organisational resources, training structure, and wider determinants of health in supporting MECC delivery, alongside skills in rapport-building. Non-trained staff noted gaps in MECC awareness, recording systems, and training but recognised MECC importance and impact. Both groups identified time constraints, confidence, and the integration of MECC within their professional roles as key factors influencing delivery of weight-related conversations.

**Conclusion:** MECC training positively impacted healthcare professionals’ perceptions and confidence in delivering weight-related MECC conversations in mental health settings. While only confidence changes were evident in surveys, qualitative findings showed multifaceted influences on MECC implementation, emphasising the need for sustained support and system-level changes to overcome structural barriers and ensure long-term MECC delivery.

## Introduction

Making Every Contact Count (MECC) is a public health strategy designed to support health behaviour change through opportunistic, person-centred conversations during routine interactions [1]. Underpinned by behavioural science, MECC applies evidence-based behaviour change techniques to address topics such as smoking, physical activity, healthy eating, alcohol, and mental health [2]. While typically delivered by health and social care professionals, MECC conversations can occur in various settings and involve anyone [1]. Conversations range from very brief interventions, such as signposting, to more in-depth discussions, depending on the needs and opportunities presented. The MECC approach has evolved to include mental health and the wider determinants of health under the term MECC Plus [2].

Healthcare professionals recognise the importance of incorporating opportunistic health and wellbeing conversations into routine care to support behaviour change, but the delivery of these interventions often falls short due to systemic and contextual barriers, including time constraints during appointments and competing priorities [3-4]. Research highlights organisational goals, leadership support, and availability of resources have influenced the adoption and implementation of MECC in Northeast England [5]. Healthcare professionals within community-based organisations acknowledge the potential of health and wellbeing conversations to address health inequalities but express apprehension about their capacity to deliver these effectively due to concerns about damaging relationships or exacerbating issues [6]. These findings highlight the need for a coordinated infrastructure to support healthcare professionals in embedding MECC into practice and overcoming barriers to delivery.

MECC training has been shown to positively impact healthcare professionals, enhancing their ability to support behaviour change and implement MECC more effectively than non-trained staff [7]. Although healthcare professionals recognise MECC as a valuable tool for behaviour change, recent findings suggest an inconsistent approach to training across organisations, highlighting the need for a more streamlined training framework [8]. To address this, online training programmes have been developed, increasing staff knowledge of behaviour change techniques [9-10]. Post-training evaluations demonstrate significant improvements in behavioural determinants, such as self-efficacy, attitudes, and outcome expectancies, indicating that MECC training enhances staff competence in behaviour change support [9]. Further research by Haighton et al. [3] emphasised the importance of integrating theoretically relevant behaviour change techniques to strengthen MECC implementation. Evaluations using the Theoretical Domains Framework (TDF) have shown increased staff confidence and goal clarity following training, with these improvements maintained at 6-10⍰weeks follow-up [10].

Across Northeast England, a bespoke training programme has been developed to include Core MECC training with the addition of A Weight Off Your Mind (AWOYM). AWOYM is a regional weight management plan developed in Northeast England to support people living with learning disabilities or severe mental illness (SMI) in managing their weight (https://www.cntw.nhs.uk/services/a-weight-off-your-mind/). AWOYM focuses on Physical Activity and Diet to help service users in inpatient mental health settings achieve a healthy weight, which can be facilitated through the delivery of weight-related MECC conversations. The bespoke MECC training adopts the ‘3As approach’ to brief interventions (‘Ask’; ‘Assist’; ‘Act’) which is adapted from the 5 As approach (‘Ask’, ‘Assess’, ‘Advise’, ‘Agree’, ‘Assist’) [11] and shortened to adhere to the brief element of MECC.

This study aimed to explore the perceptions of delivering weight-related MECC conversations using the COM-B framework, which identifies Capability, Opportunity, and Motivation as key drivers of behaviour change [12]. For example, this includes whether staff feel confident (capability), have time and resources (opportunity), and believe MECC is worthwhile (motivation). The study assessed perceived capabilities, opportunities, motivation, and experiences of staff in a mental health inpatient setting delivering weight-related MECC conversations to service users.

## Method

### Design

This study employed a mixed-methods design, combining quantitative and qualitative approaches to explore staff’ experiences and perspectives of MECC delivery. Quantitative data were collected through a structured survey, providing an overview of key patterns of capabilities, opportunities and motivation, while qualitative data were gathered through semi-structured, one-to-one interviews to generate in-depth views. The qualitative approach was guided by the Theoretical Domains Framework (TDF), which facilitated a systematic exploration of factors influencing participants’ behaviours and perspectives. Ethical approval for this study was granted from Northumbria University Faculty of Health and Life Sciences Ethics Committee (ref 43,190). The study and protocol were approved by the R&D department at St Nicholas Hospital, Cumbria, Northumberland, Tyne & Wear NHS Foundation Trust. The protocol was published [13] and pre-registered on the Open Science Framework: osf.io/ewktc. This study and related findings are reported in line with the guidance for mixed methods studies from the EQUATOR Network website [14].

### Professional contributor panel

A group of healthcare professionals volunteered to provide feedback on survey and interview design and delivery. Three female healthcare professionals from a healthcare organisation in Northeast England participated in regular online meetings via Microsoft Teams. Their feedback helped improve elements such as survey layout, question format, and interview topic guides.

### Setting and participants

This project was conducted in an NHS mental health inpatient setting in Northeast England. Study participants included both inpatient and outpatient staff, comprising healthcare professionals caring for patients with severe mental illness, including those in secure services.

### Data collection and analysis

All participants received a participant information sheet and provided informed consent prior to participation, and were assured that all data would be anonymised before a study debrief.

#### Quantitative online staff survey

A quantitative cross-sectional online survey was used to capture staff perspectives on the implementation of MECC following either Train the Trainer (TtT) or Core MECC+AWOYM training. Surveys were distributed at three time points: pre-training (immediately prior), post-training (immediately after), and follow-up (8–10 weeks later). Between September 2022 and July 2023, all trained staff (N=52) were invited to participate.

Convenience sampling was used to recruit staff working in mental health inpatient setting for the survey. The research team collaborated with a Health Improvement Specialist (SF) to distribute surveys internally, ensuring confidentiality. Surveys and interview sign-up forms were created in Qualtrics, with links and QR codes shared via email and during training sessions. Pre-training surveys were completed at the start of each session, post-training surveys at the end, and follow-up surveys via email. Each survey took approximately 5–7 minutes to complete, and participants generated unique codes for dataset identification.

The surveys (Supplementary file 1), tailored to pre-training, post-training, and follow-up, incorporated validated scales relating to COM-B (Capability, Opportunity, Motivation and Behaviour) [4], items adapted from ‘Making MECC work’ [15] and items assessing ‘Confidence, Usefulness and Importance’ of delivering MECC [10].

To account for incomplete and unmatched samples across the three data collection points (T1, T2, T3), both within-subject and between-subject approaches were used. First, a subset of matched participants (n = 15 for T1–T2) was analysed using paired-samples t-tests to explore within-subject changes over time. Given that only four participants provided matched responses across all three time points, a repeated measures ANOVA was not considered appropriate due to insufficient statistical power. For the larger dataset, where samples at each time point were partially or fully unmatched, responses were treated as independent. Independent-samples t-tests were then conducted to compare mean scores between two time points (e.g., T1 vs T2, T2 vs T3). Given the relatively small sample sizes, we also ran equivalent non-parametric tests (Wilcoxon signed-rank and Mann–Whitney U) to check the robustness of results. Parametric tests (t-tests) are presented for clarity and interpretability; equivalent non-parametric tests produced consistent results. Analyses used available-case data, and sample sizes vary across tests due to missing responses. Valid numbers for each comparison is reported in the results. All analyses were conducted using IBM SPSS Statistics (version 26).

#### Qualitative staff interviews

Purposive sampling was used to recruit healthcare professionals across inpatient and community-based settings, ensuring maximum variation in professional grouping, age, and gender. Both trained and non-trained staff were included to understand barriers, knowledge gaps, and perspectives from those with and without MECC training, which is crucial for identifying needs and tailoring effective training programs. Comparing trained and non-trained perspectives also highlights the impact of training and areas for improvement. Healthcare professionals who opted in and provided contact details through the sign-up link were then contacted by a member of the research team (EK) to schedule an interview.

A qualitative interview study was conducted with staff who had received Train the Trainer, Core MECC + A Weight Off Your Mind, or no MECC training. The aim was to understand their views on delivering MECC as either trained or non-trained staff.

Interviews were conducted online via Microsoft Teams between May 2023-November 2023, by an experienced qualitative researcher (EK, Female) and lasted 20-45 minutes. A topic guide (Supplementary file 2) was informed by previous work in this area [5, 16] and developed based on feedback from professional contributors, and initial participant interactions. The guide focused on the acceptability of bespoke MECC training compared to generic behaviour change training, and key enablers and barriers to implementing MECC in mental health settings. For interviews with non-trained staff, examples of MECC conversation starters (Supplementary file 2) were presented to explore how they might be used in patient interactions. Data collection was guided by the concept of information power [17] and continued until the research team determined that the interviews had gathered sufficient information to address the study’s objectives. We initially estimated 30 interviews (approximately 15 per group), with final sample size determined through iterative analysis. Author EK reviewed the transcripts for accuracy and ensured they were anonymised.

Interviews were recorded, transcribed verbatim and anonymised. Transcripts were analysed using framework method [18] on NVivo V.12. An initial coding framework was developed collaboratively by two researchers (EK and AR) through independent coding of 10% of transcripts. This framework was applied to all transcripts by EK. We employed an inductive thematic analysis method [19], allowing themes to be generated from the data, a common practice in studies guided by the Theoretical Domains Framework (TDF) [20].

## Results

### Quantitative online staff survey

Survey data were analysed from staff who completed the pre, post, and follow-up surveys. Table 1 shows the characteristics of 42 participants who responded to the surveys (note that response rates varied). Most participants were women in nursing roles within inpatient settings, primarily attending Train the Trainer training. The largest age groups were 25-34 and 45-54 (26.1% each), followed by 35-44 (23.8%). The majority had over 5 years’ experience (40.5%) in their role, with 26.2% having 6 months’ or less.

#### Effects on COM-B items, confidence, usefulness, and importance of MECC delivery

A total of 49 participants completed the variables of interest at pre-test (T1), 27 completed the post-test (T2), and 10 responded at follow-up (T3). A subset of 15 participants provided matched data at both T1 and T2, while only 4 participants completed all three time points (T1, T2, and T3). Due to missing data and incomplete matching across time points, sample sizes vary between analyses. All reported sample sizes in Table 2 and 3 refer to the number of valid cases included in each specific test. As a result of participant attrition, analyses were conducted using both matched and independent samples where appropriate.

**Table 1.** Survey participant characteristics.

| Characteristic (n=number of responses) | n(%) |
| --- | --- |
| **Gender (n=42)** |  |
| Male | 9 (21.4) |
| Female | 32(76.2) |
| Non binary/ Third gender | 1(2.4) |
| **Age (n=42)** |  |
| 18-25 | 2(4.8) |
| 25-34 | 11(26.1) |
| 35-44 | 10(23.8) |
| 45-54 | 11(26.1) |
| 55+ | 8(19.1) |
| **Current role (n=42)** |  |
| Nurse/Nurse associate | 21(50) |
| Allied healthcare professionals | 4(9.5) |
| Healthcare Assistant | 6(14.3) |
| Other (Including Tobacco treatment advisor, Peer support worker and trainee/assistant psychologist) | 11(26.2) |
| **Time in role (n=42)** |  |
| 6 months or less | 11(26.2) |
| Over 6 months, up to 1 year | 6(14.3) |

| **Characteristic (n=number of responses)** | **n(%)** |
| --- | --- |
| Over 1 year, up to 3 years | 6(14.3) |
| Over 3 years, up to 5 years | 2(4.8) |
| Over 5 years | 17(40.5) |
| **Ward/Service location (n=42)** |  |
| Inpatient | 25 (59.5) |
| Community | 17(40.5) |
| **Training received (n=34)** |  |
| Train the Trainer | 22(52.4) |
| Core MECC+AWOYM | 12 (28.6) |
| **Delivery method (n=34)** |  |
| Online | 23(54.8) |
| Face to face | 11(26.2) |

**Table 2.** Mean and standard deviation and key comparisons across time points for COM-B items, confidence, importance and usefulness outcomes (paired t-test statistics T1 vs T2).

| Outcome | T1 Mean (SD) | T2 Mean (SD) | n | Paired t-test statistics T1 vs T2<sup>1</sup> |
| --- | --- | --- | --- | --- |
| Physical Capability | 8.1 (1.9) | 8.3 (1.9) | 15 | $t(14) = -.41, p = .69, d = -.11$ |
| Psychological Capability | 8.3 (2.0) | 8.0 (1.8) | 15 | $t(14) = .70, p = .49, d = 0.18$ |
| Physical Opportunity | 66.7 (28.2) | 71.8 (23.1) | 14 | $t(13) = -.77, p = .46, d = -0.21$ |
| Social Opportunity | 67.0 (20.3) | 67.4 (27.3) | 10 | $t(9) = -.04, p = .97, d = -0.01$ |
| Automatic Motivation | 7.9 (2.1) | 8.3 (2.0) | 15 | $t(14) = -.57, p = .58, d = -0.15$ |
| Reflective Motivation | 9.6 (0.7) | 8.8 (2.3) | 15 | $t(14) = 1.25, p = .23, d = 0.32$ |
| Confidence | 7.4 (2.1) | 8.4 (1.7) | 14 | $t(13) = -1.36, p = .20, d = -0.36$ |
| Importance | 9.1 (1.2) | 8.9 (1.5) | 14 | $t(13) = .49, p = .64, d = 0.13$ |
| Usefulness | 8.6 (1.8) | 7.9 (1.8) | 14 | $t(13) = 1.33, p = .21, d = 0.35$ |

<sup>1</sup> Paired-sample t-test based on matched participants for T1 and T2. Sample sizes reflect valid cases per comparison. Cohen's *d* effect sizes interpreted as small ( $d \approx .2$ ), medium ( $d \approx .5$ ), or large ( $d \approx .8$ ).

**Table 3.** Mean and standard deviation and key comparisons across time points for COM-B items, confidence, importance and usefulness outcomes (independent t-test statistics).

| Outcome | T1 Mean (SD, n) | T2 Mean (SD, n) | T3 Mean (SD, n) | Independent t-test statistics T1 vs T2 | Independent t-test statistics T2 vs T3 |
| --- | --- | --- | --- | --- | --- |
| Physical Capability | 8.0 (2.1) (n=48) | 8.3 (2.1) (n=27) | 8.7 (1.7) (n=10) | T1 vs T2: $p = .60, t(73) = -.53, d = -0.15$ | T2 vs T3: $p = .31, t(35) = -.50, d = -0.18$ |
| Psychological Capability | 8.0 (2.1) (n=48) | 8.3 (1.9) (n=27) | 8.4 (2.4) (n=10) | T1 vs T2: $p = .53, t(73) = -.63, d = -0.13$ | T2 vs T3: $p = .85, t(35) = -.19, d = -0.07$ |
| Physical Opportunity | 63.5 (31.3) (n=39) | 69.0 (26.0) (n=27) | 49.2 (29.2) (n=9) | T1 vs T2: $p = .46, t(64) = -.75, d = -0.19$ | T2 vs T3: $p = .06, t(34) = .63, d = 0.74$ |
| Social Opportunity | 61.7 (25.2) (n=30) | 65.0 (27.2) (n=27) | 33.0 (32.1) (n=10) | T1 vs T2: $p = .64, t(55) = -.47, d = -0.12$ | T2 vs T3*: $p = .01, t(35) = .61, d = 1.12$ |
| Automatic Motivation | 7.5 (2.1) (n=48) | 8.3 (1.9) (n=27) | 8.2 (1.9) (n=10) | T1 vs T2: $p = .12, t(73) = -1.6; d = -0.38$ | T2 vs T3: $p = .89, t(35) = .13, d = 0.05$ |
| Reflective Motivation | 8.9 (2.0) (n=49) | 8.7 (2.0) (n=27) | 8.7 (1.5) (n=10) | T1 vs T2: $p = .72, t(74) = .36, d =$ | T2 vs T3: $p = .99, t(35) = .01,$ |
| | | | | 0.09 | $d = 0.0$ |
| Confidence | 7.5 (1.8)<br>(n=47) | 8.2 (1.8)<br>(n=27) | 8.5 (1.2)<br>(n=10) | T1 vs T2: $p = .09$,<br>$t(72) = -1.7$, $d = -0.41$ | T2 vs T3: $p = .33$, $t(35) = -.44$, $d = -0.16$ |
| Importance | 8.9 (1.3)<br>(n=47) | 9.0 (1.5)<br>(n=27) | 8.5 (1.6)<br>(n=10) | T1 vs T2: $p = .71$,<br>$t(72) = -.37$, $d = -0.09$ | T2 vs T3: $p = .17$, $t(35) = .98$,<br>$d = 0.36$ |
| Usefulness | 8.0 (1.9)<br>(n=47) | 7.8 (1.8)<br>(n=27) | 6.2 (7.0)<br>(n=10) | T1 vs T2: $p = .56$,<br>$t(72) = .58$, $d = 0.14$ | T2 vs T3: $p = .50$, $t(35) = .70$,<br>$d = 0.41$ |

Note: Sample sizes reflect valid cases per comparison. Cohen's $d$ effect sizes interpreted as small ( $d \approx .2$ ), medium ( $d \approx .5$ ), or large ( $d \approx .8$ ). \* $p < .05$ , $p$ -values are two-tailed unless otherwise specified.

As shown in Table 2, paired-samples t-tests conducted on the matched subset of participants (n = 15) to examine within-person change from pre-test (T1) to post-test (T2) revealed no significant differences across any of the measured outcomes (all p > .10). Non-parametric analyses yielded the same overall pattern of results.

To include all available data, two independent-samples t-test were also conducted using the full pre-test (T1, n = 49), post-test (T2, n = 27), and follow-up groups (T3, n = 10). Analyses revealed a statistically significant increase in confidence from T1 to T2 in the expected direction, as indicated by a one-tailed independent t-test (p = .046); however, this effect did not reach significance using a two-tailed criterion (p = .09). Between T2 and T3, a significant decrease in perceived social opportunity was observed (p = .02), while perceived physical opportunity also declined but did not reach statistical significance (p = .06). Results from non-parametric analyses were consistent with the parametric tests, with the exception of usefulness, where the Mann–Whitney U indicated a significant decrease over time (U = 33, p < .001), consistent with the direction of the t-test findings. Please see Table 3 for full details.

### Qualitative staff interviews

Out of the 25 individuals who participated in the interviews, 10 had received no prior MECC training and 15 had received MECC training which included either Core MECC with A Weight off Your Mind, or Train the Trainer. The individuals represented those working in management and leadership roles (n=5) with little direct contact with service users, to frontline staff such as nurses (n=10), peer support workers, healthcare assistants, sport and exercise therapists and other health-related roles (n=10) with daily or intermittent contact with service users Table 3 provides an overview of participant characteristics of staff who took part in interviews.

**Table 3.** Interview participant characteristics.

| Participant | Trained/Non trained | Role | Time in role | Age |
| --- | --- | --- | --- | --- |
| Male | Non trained | Medic | 5years+ | 55+ |
| Female | Non-trained | Healthcare assistant | <6months | 25-34 |
| Female | Non-trained | AHP-OT | 5 years+ | 45-54 |
| Male | Non-trained | Nurse | 5 years+ | 55+ |
| Female | Non-trained | Nurse | 5 years+ | 55+ |
| Female | Non-trained | Student- mental health nursing | 1-3 years | 35-44 |
| Female | Non-trained | Admin lead | 5 years+ | 35-44 |
| Female | Non-trained | Nurse | 5 years+ | 55+ |
| Female | Non-trained | Associate director for involvement and lived experience | 5 years+ | 55+ |
| Female | Non-trained | Assistant psychologist | 1-3 years | 25-34 |
| Female | Trained (CM+AWOYM) | Nurse | 5 years+ | 45-54 |
| Female | Trained | Nurse | 5 years+ | 45-54 |
|  | (CM+AWOYM) |  |  |  |
| **Female** | Trained<br>(CM+AWOYM) | Head of<br>workforce and<br>OD | 5 years+ | 55+ |
| **Female** | Trained (TtT) | Nurse | 5 years+ | 35-44 |
| **Female** | Trained<br>(CM+AWOYM) | Nurse | 5 years+ | 35-44 |
| **Female** | Trained<br>(CM+AWOYM) | Nurse | 5 years+ | 45-54 |
| **Female** | Trained<br>(CM+AWOYM) | Pharmacy | 3-5 years | 25-34 |
| **Male** | Trained<br>(CM+AWOYM) | Sport/Exercise<br>therapist | 1-3 years | 25-34 |
| **Male** | Trained<br>(CM+AWOYM) | Sport/Exercise<br>therapist | 5 years+ | 35-44 |
| **Female** | Trained (TtT) | Tobacco<br>treatment<br>advisor | 6 months-1 year | 55+ |
| **Female** | Trained<br>(CM+AWOYM) | Peer Supporter<br>Supervisor | 5 years+ | 25-34 |
| **Female** | Trained<br>(CM+AWOYM) | Nurse | 1-3 years | 45-54 |
| **Male** | Trained (TtT) | Mental Health<br>case manager | 5 years+ | 45-54 |
| **Female** | Trained (TtT) | Healthcare<br>assistant | 5 years+ | 45-54 |
| **Female** | Trained (TtT) | Peer Support | 6 months- 1 year | 35-44 |

## Main results

Analysis found nine domains of influence for delivering weight-related conversations across trained and non-trained staff interviews. Several sub-themes were created based on each domain (Table 4). Trained staff emphasised the significance of organisational resources, training structure, and wider determinants of health in supporting MECC delivery, alongside skills in rapport building and an attuned, empathic approach to emotional needs. Non-trained staff highlighted gaps in MECC awareness, recording systems, and necessary training but recognised its importance and impact on patient care. Both groups identified time constraints, confidence levels, and the integration of MECC within their professional roles as key factors influencing effective implementation.

**Table 4.** Factors influencing weight-related MECC conversations, including themes, sub-themes, and illustrative quotes, mapped to TDF domains.

| **TDF Domain (COM-B element)** | **Trained staff (Sub-theme)** | **Non-trained staff (Sub-theme)** |
| --- | --- | --- |
| **Environmental Context** | Organisational | Organisational |
| **(Physical Opportunity)** | resources<br>• Organisation of training<br>• Wider determinants of health<br>• Time constraints | resources<br>• Awareness of MECC and promoting training<br>• Wider determinants of health<br>• MECC recording systems<br>• Time constraints |
| **Beliefs about Consequences (Reflective Motivation)** | • Benefits to patients<br>• Therapeutic relationship<br>• Training outcomes<br>• Organisational benefits | • Importance of MECC<br>• Embedding MECC<br>• Impact on patients |
| **Skills (Psychological Capability)** | • Knowing when to use MECC<br>• Delivering weight-related MECC<br>• Building a rapport with patients<br>• Emotional skills needed to deliver MECC to patients with SMI<br>• Applying skills acquired from training into practice | • Knowing when to deliver MECC<br>• Delivering MECC<br>• Build rapport<br>• Previous training |
| **Knowledge (Psychological Capability)** | • Training content<br>• What MECC entails<br>• Information suitable for patient | • Lack of awareness of MECC<br>• What MECC entails<br>• What training needed |
| **Beliefs about Capabilities (Reflective Motivation)** | • Confidence to deliver MECC | • Confidence to use MECC |
|  | • Therapeutic relationship | • Training useful for role |
| **Professional role (Reflective Motivation)** | • MECC as part of role<br>• Contact with patients | • How MECC fits with role<br>• Contact with patients |
| **Behavioural regulation (Psychological Capability)** | • Nature of the behaviour (initiating MECC conversations)<br>• Planning and feedback | • Nature of behaviour<br>• Action planning |
| **Intention and goals (Reflective Motivation)** | • Motivation to deliver MECC<br>• Patient motivation | • Intention to deliver MECC |
| **Social influences (Social Opportunity)** | • Cultural factors of MECC conversations<br>• Providing social support to patients | • Providing social support to patients |

### Environmental context and resources *(Physical Opportunity)*

Differences emerged between trained and non-trained staff in how they accessed resources, training promotion, and addressing external challenges to delivering MECC.

#### Organisational resources

Trained staff, familiar with tools like the MECC Gateway website (https://www.meccgateway.co.uk/nenc), accessed various resources to support MECC conversations. Whereas non-trained staff reported limited access to resources, which added pressure to their role.

> *‘Resources as well. Services we use outside of NHS that receives social care, care packages, all that kind of stuff, that puts a…that being not available puts a pressure on our role.’ Participant 9, non-trained*

#### Awareness and promoting training

Trained staff noted organisational awareness and direct invitations from management to attend training. Non-trained staff highlighted having an awareness of training such as being informed via advertisement in staff bulletins, however, did not appear to have been approached directly from management to take part in any MECC related training.

> *‘I think I have seen stuff generally advertised on the intranet of our email about MECC training. I haven’t been approached as an individual about MECC training.’ Participant 3, non-trained*

#### Wider determinants of health

Both trained and non-trained staff identified socioeconomic barriers, such as affordability of healthy food, as challenges to promoting healthy lifestyle changes:

> *‘I mean, we’re talking about a high deprivation area. We’re talking about people that don’t necessarily have a lot of money for what we would think is the right things.… You know, and if you want your mental health to get better then maybe we need to have some structure during the day as to what you’re occupying your time with.’ Participant 17, trained*

#### Time constraints

Limited time during consultations and high caseloads hindered MECC delivery. Staff noted that documenting MECC conversations added to this challenge:

> *‘Often we’ve got somebody coming into the room straight after, so you can’t run over so like if you’ve a got certain number of things that you need to get done in that hour obviously adding on extra stuff like delivery of sort of brief interventions, MECC etc. you’re going to struggle because then you’re going to impact on the next person’s time as well.’ Participant 23, trained*

#### MECC recording systems

Both trained and non-trained staff discussed the recording of MECC conversations and reported a lack of standardised recording systems for MECC, often documenting conversations as wellbeing discussions instead. Also, the electronic patient record system was used for documenting patient contact but lacked a clear category for MECC.

> *‘So, I don’t, it’s not titled a MECC conversation. It’s kind of classed under wellbeing discussion.’ Participant 19, trained*

### Beliefs about consequences *(Reflective Motivation)*

This domain reflected how both trained and non-trained staff perceived the delivery of MECC, highlighting benefits for patients, the importance of embedding MECC in practice, and its potential to enhance therapeutic relationships and organisational outcomes.

#### Benefits to Patients

Trained staff believed that MECC conversations could improve patient health and wellbeing, providing a framework for support if patients were receptive to treatment. As one trained staff member shared:

> *‘Hopefully make small changes and help the patients live a happier and healthier lifestyle I would say.’ Participant 14, trained*

Non-trained staff also acknowledged the potential impact, noting that while they were already having similar conversations, the absence of formal training limited their ability to implement MECC effectively. A non-trained staff member explained:

> *‘Knowing that they might want to get involved or they might not want to, erm I’d say if it’s a part of the intervention and it’s a part of the care plan then they’d be more - they’ll be more inclined to just want to help us help themselves essentially.’ Participant 6, non-trained*

#### Therapeutic relationship

Trained staff noted that MECC conversations helped build stronger therapeutic relationships by offering a chance to better understand patients’ health and wellbeing. One participant said:

> *‘Because it’s pivotal to getting to know your patient. It’s kind of like finding out whereabouts they are with their health.’ Participant 17, trained*

Additionally, these conversations encouraged continuity of care, with the view that patients will remember advice from past interactions, resulting in more tailored care and signposting to suitable information and services:

> *‘They’ll remember that you took that time to listen to them, they’ll remember that you were the one who suggested, you know, they did this or whatever and so I think that really helps.’ Participant 23, trained*

Non-trained staff members highlighted a concern that raising the topic of weight management with people living with overweight could be detrimental and push patients away:

*‘I guess my concern would be it’s about people who are overweight will have had those conversations with other people. And so, it’s sort of gauging that, isn’t it? Because, what I would hate to do is push people away’. Participant 1, non-trained*

#### Organisational benefits

Both trained and non-trained staff identified organisational benefits of MECC, particularly by spreading more awareness of MECC through training, this can lead to MECC being embedded in the trust and can have positive outcomes including better communication between services through signposting and referrals:

> *‘Usually you’ll speak about smoking and not say like oh, I don’t want to smoke any more and you go oh right, well we’ve got the Quit team, that’s another thing in place that enables MECC.’ Participant 14, trained*

Non-trained staff suggested that making MECC training mandatory could ensure wider adoption across the trust:

> *‘I think it should be mandatory training, certainly. Then it’s something that everybody is going to have an awareness of. I think the more people are aware, the more they’re going to build it into their practice, and that can only be a good thing’. Participant 2, non-trained*

### Skills *(Psychological Capability)*

Both trained and non-trained staff highlighted various skills critical to delivering MECC conversations, including recognising appropriate moments for intervention, building rapport with patients, and applying knowledge acquired through training.

#### Knowing when to use MECC

Staff emphasised understanding patient cues to determine when MECC conversations are appropriate, often preferring to intervene later in a patient’s recovery journey:

> *‘I think you’ve just got to be led by the person when you’re in discussion with them. I think you have to read them, and obviously, be quite tactful.’ Participant 4, non-trained*

> *‘Because we do deal with some really unwell people, and when they first come in, obviously, that’s not the right time to have the conversation. And I find it’s easier to have the discussions when they’re on their way out of the hospital, on the discharge planning side of things.’ Participant 11, trained*

#### Delivering weight-related MECC

Trained staff demonstrated confidence in initiating MECC conversations, often sparking patient interest and signposting them to resources. In contrast, non-trained staff relied on routine opportunities, such as physical health checks, for brief lifestyle discussions but lacked alignment with MECC principles. Trained staff acknowledged patient resistance but approached MECC systematically, while non-trained staff struggled with sensitive topics, fearing patients might feel judged due to limited training:

> *‘I think sometimes how we approach it, and like you said before, and like I mentioned about the first questions about that feeling of I’m judging you about the things that you do, and actually oh don’t start talking to me about healthy diet or whatever, you know.’ Participant 7, non-trained*

#### Building rapport

Both trained and non-trained staff recognised the importance of building rapport with patients, emphasising that relationships take time to develop. However, trained staff utilised their relationships to identify and address specific barriers, tailoring MECC conversations to patient needs. Non-trained staff also valued rapport but focused more broadly on engaging patients over time, lacking the targeted approach emphasised by trained staff:

> *‘Whereas doing it in a more informal way, and having that relationship with the patient as well, and looking at their barriers and stuff like that and being aware of that and trying to remove as many barriers as you can and just being really.’ Participant 22, trained*
>
> *‘I think it’s building that relationship initially and getting somebody to engage and want to work with you and talk openly with you about things, and that can sometimes take a few weeks.’ Participant 8, non-trained*

### Emotional skills needed to deliver MECC to patients with SMI

Although MECC was discussed as being opportunistic, emotional factors could affect whether a MECC conversation can occur. From a staff perspective if they did not feel ready to have a MECC conversation with a patient this would prevent the conversation occurring however, this also appeared to be in the best interest of the patient particularly if staff could envisage a long-term healthcare plan with the patient.

> *‘And the same with yourself if you’re not in the best frame of mind or whatever, it’s best to make sure that you’re ready to have that conversation as well because it can end up being a long-term thing.’ Participant 22, trained*

Having the skills to understand the emotional state of the patient also influenced if staff could initiate a MECC conversation. Patients who were highly emotional or in crisis may not benefit from MECC conversations as a different intervention might be more suitable or patients may not be willing to engage in MECC advice if they are feeling low or emotional.

> *‘Again, it varies. If they’re quite emotional, if they’re quite down it might not be the right time and a lot of time they come in in crisis or something that they deem is a crisis and they just need to talk it through, so you get sidelined.’ Participant 17, trained*

#### Applied skills acquired from training into practice

Trained staff emphasised that MECC training directly enhanced their practical skills for initiating and conducting MECC conversations. These skills included knowing how to approach patients, use conversation starters, and confidently signpost resources. Training provided clear examples and strategies, which helped staff feel more equipped to deliver MECC conversations effectively:

> *‘So, find out what they-so, what I’ll do is I’ll start a conversation with the service users about whichever kind of topic it is. And then, I’ll be like, “Look, I’ll go away. I’ll do a bit of research for you. Find some stuff for you to look at, if this is what you’re interested in.’ Participant 11, trained*

### Knowledge *(Psychological Capability)*

This subtheme highlights the pivotal role of MECC training in enhancing staff knowledge and understanding of how to implement MECC conversations effectively. Both trained and non-trained staff discussed the importance of being well-informed, but key differences emerged in how knowledge was gained and applied.

#### Training content

Trained staff particularly valued the practical aspects of the MECC training, including how to signpost patients to relevant resources. They appreciated the guidance on where to find resources and how to use them during MECC conversations.

> *‘But I think it’s mainly the resources and things like that, and how to start the conversations that I wanted to learn. I wanted to learn because obviously, I don’t know everything. And being able to signpost people to things that are really useful for them, was really useful for me.’ Participant 11, trained*

Trained staff also noted the training’s emphasis on patient-tailored care, which helped them understand the best times to introduce MECC conversations and provide brief advice.

> *‘It did emphasis a lot on patient tailored care, kind of thing, so like we were saying before, like it might not necessarily be the exact space to take, to have this conversation with a patient.’ Participant 14, trained*

#### What MECC entails

A solid knowledge base of MECC, often gained through training, helped staff confidently engage with patients about health topics. Trained staff emphasized that the evidence and background knowledge learned in training allowed them to approach MECC conversations more effectively. While aware of health promotion, non-trained staff lacked the deeper understanding of the structured MECC model. They still recognised its value but had less clarity on how to implement it systematically:

> *‘I would say I know that it’s about making every contact count and just about how making opportunities in everyday simple conversations about promoting health and wellbeing, health promotion and all aspects of that, and that can be a simple check in from checking someone’s had a drink or they’ve been to the GP or how they’re physically feeling, so it’s just trying to interweave those conversations into your everyday practice.’ Participant 7, non-trained*

#### Lack of awareness of MECC

Staff who had not received MECC training exhibited limited knowledge of the model and its implementation. Many were unaware of the specifics of MECC, which hindered their ability to use it effectively:

> *‘I don’t really know much about it other than I guess what the title kind of implies, but I don’t really know the ins and outs of it.’ Participant 9, non-trained*

#### Information suitable for patient

Both trained and non-trained staff discussed the challenge of providing information that patients could easily understand. Non-trained staff mentioned giving general health promotion. In contrast, trained staff took additional steps to ensure resources were accessible, often seeking out easy-read formats or explaining complex materials to ensure patients comprehended the information.

> *‘The development of social stories or pictorial information around that would help, so if there was every source around easy read information that the nurse could maybes use or even adapt to the individual as well, you know. So, pictures and things that you could adapt to help that person understand, perhaps would be helpful’. Participant 20, trained*.

### Beliefs about capabilities *(Reflective Motivation)*

Staff discussed confidence and capability in delivering MECC. Training enhanced confidence among trained staff, while non-trained staff felt training would improve their ability.

#### Confidence to deliver MECC

Trained staff gained confidence from training and frequent practice, particularly in accessing resources and understanding patient reasoning:

> *‘I feel like my confidence has grown quite a bit. I’ve learned how to say, “I’ll find out that information for you.” And walk away from it feeling confident that I can find the information. I’m a lot more confident in starting the conversations, and a lot more confident in having those discussions with people about the MECC training. Well, not the training itself, but the conversations’. Participant 11, trained*

Non-trained staff, though less confident, still initiated lifestyle conversations and noted that building rapport eased MECC discussions:

> *‘I wouldn’t say I’m overly confident, but it’s not an issue having the conversations. But it is a learning curve. I’m still I’ve only been doing the job about a year so I’m still building up relationships with the staff and as that’s going it is getting easier and much more comfortable to do.’ Participant 5, non-trained*

#### Therapeutic relationship

Strong patient relationships facilitated MECC conversations by creating opportunities to explore health and wellbeing topics and offer advice:

> *‘We’ll just lean on our therapeutic relationship with patients to be able to, so just to add that extra confidence that if they are struggling with something they’re going to bring it up themselves or we can coax it out of them by just having a little bit of banter.’ Participant 18, trained*

#### Training as an enabler

Non-trained staff emphasised the importance of MECC training for improving conversations and implementing MECC effectively:

> *‘But I feel like if staff had the training for it there’d be a lot more like implementation of those kinds of discussions.’ Participant 6, non-trained*

Half of the non-trained staff members also mentioned that taking part in Train the Trainer training would be useful for their role and allow them to cascade training to colleagues or smaller staff teams which would help with implementation of MECC across the trust, however some mentioned having the time to deliver cascade training would be an issue:

> *‘So, I’d prefer to do the Train the Trainer one because I thought it would be useful within my role.’ Participant 7, non-trained*

### Professional role *(Reflective Motivation)*

Both trained and non-trained staff recognised that MECC is an integral part of their professional role, especially in roles with limited patient contact. However, differences emerged in how MECC was applied depending on training and the nature of staff roles.

#### MECC as part of role

Both trained and non-trained staff agreed that MECC aligns with their responsibilities. Non-trained staff emphasised how MECC supports broader recovery and rehabilitation efforts, focusing on helping patients return to their communities.

> *‘Our job is, like I said, recovery and rehabilitation, so we look at all of the aspects that people might need to attend to, to be able to get themselves back on their feet, to be able to get back out there in the community and live. A lot of that is to do with just these sorts of issues, so they’re fundamental to what we do.’ Participant 2, non-trained*

#### Contact with patients

Staff in roles with less patient contact highlighted that MECC’s brief advice model is well-suited to their roles, where opportunities for ongoing patient engagement are limited. The flexibility of delivering MECC in short interactions made it more feasible for these staff to incorporate into their daily tasks. In contrast, staff with more scheduled or ongoing contact with patients recognized that while MECC could be integrated into consultations, they often lacked the opportunity to follow up on advice once their contact with the patient concluded. This made it difficult to monitor the long-term impact of their brief MECC interventions.

> *‘So, we don’t have, you know, see this patient for six appointments or something like that, and then that would be me that would be finished, so it could be that six appointments you might finish that contact with that person.’ Participant 10, non-trained*

### Behavioural regulation *(Psychological Capability)*

While both trained and non-trained staff face similar barriers to delivering MECC, trained staff tended to have a more structured approach, benefiting from additional planning, knowledge, and feedback, which enhanced their ability to engage patients effectively. Non-trained staff, however, felt less supported and often lacked the preparation to confidently initiate and follow through with MECC conversations.

#### Nature of the behaviour (Initiating MECC conversations)

Both trained and non-trained staff mentioned offering advice on healthy eating, with non-trained staff focusing on providing referrals and basic diet education.

> *‘But also, erm if they’re wanting to change their eating habits and that they also need to be referred to a dietician or just probably like them in general we could give them some education around that as well. Like erm what’s the best foods to eat, diet and exercise and things like that, so yeah.’ Participant 9, non-trained*

A key barrier to delivering MECC conversations mentioned was the patient’s wellness. Trained staff noted that patients with serious mental health conditions were often too unwell to engage in MECC discussions. Non-regular contact with patients, especially in roles with less continuity, was identified as another barrier, as it hindered the development of trust and engagement in MECC conversations.

> *‘Specifically, when it comes to MECC, sometimes people are just too unwell to have that conversation. Especially working in a mental health hospital, that some people are-they don’t think they’re unwell at that time, or they don’t think it is an issue, or they’re too unwell to engage in those types of conversations.’ Participant 11, trained*

#### Planning and feedback

Staff discussed planning MECC conversations in advance, with trained staff highlighting the importance of preparation to ensure the conversation was delivered effectively and in a private setting.

> *‘So, I would kind of plan how I would bring it up before, you know, because if you don’t plan how you’re going to bring something up it can come out wrong. So, you plan how you were going to bring it up and make sure that you’re in a private conversation rather than chatting about it when everyone else is there. Or if they’re happy to, you know. Sort of people are quite happy to do that.’ Participant 22, trained*

Regarding feedback, trained staff, especially those in roles where MECC is actively monitored, mentioned receiving feedback on their conversations.

### Intention and goals *(Reflective Motivation)*

Staff discussed their motivation for delivering MECC, with trained staff showing a stronger personal interest in health promotion and a desire to share their knowledge with colleagues. They also highlighted the importance of positive reinforcement to maintain patient motivation. Non-trained staff viewed MECC training as a professional responsibility, recognizing its value in enhancing their skills. Both groups acknowledged the role of patient motivation but differed in their confidence and approach to initiating health conversations.

#### Motivation to deliver MECC

Trained staff showed strong motivation to initiate MECC conversations, often driven by a personal interest in health promotion. This interest was particularly evident in their desire to engage both patients and their families.

> *‘I just think I’m very interested in health promotion in general and I just think you’ve got people, I mean for us, for my sort of role, it’s actually more probably parents than the actual children who I work with, you know.’ Participant 23, trained*

#### Patient motivation

Trained staff emphasised the importance of feedback in maintaining patient motivation to make healthier lifestyle changes, particularly in settings where motivation could fluctuate:

> *‘So I guess the motivation, especially within secure care, can fluctuate quite a lot. It’s always quite difficult. I think feedback for patients is really important for maintaining motivation so if someone’s kind of made some change on their diet is to be fed back that and to praise them for it.’ Participant 18, trained*

Providing regular support was seen as an important part to motivating patients. Continuity of support provided to patient to maintaining the relationship meant that MECC conversation could be built upon with regular contact between healthcare professionals and patients.

> *‘Providing the support and making sure that it’s regular support and for myself not letting them down, being that reliable… at the end of every session I tell them that I’ll come and see them next week and it’ll be roughly the same time so I’m not going on a ward and they’re not there…. So there’s a motivation from them to say well, if something else came along they’ve kind of said, “No, I’ll do it then because [Name]’s coming to see me.’ Participant 12, trained*

### Social influences (*Social Opportunity*)

#### Cultural factors of MECC conversations

Trained staff highlighted that cultural factors might affect how MECC conversations are delivered, for example accessing culturally specific information which is not demonstrated throughout the training. The lack of skill and training in this area might hinder the process of providing MECC conversations to patients from different cultural backgrounds.

> *‘I think, especially, cultural factors. I do struggle with people with different cultures, for example. I wouldn’t know where to go, or things like that. That would be a massive struggle with me, I tend to find. But I think that’s more my own development and learning about stuff, that I need to look a little bit more at. I think one of the social factors that’s positive is that I’ve been there and I’ve got the T-shirt.’ Participant 11, trained*

#### Providing social support to patients

Staff discussed the importance of providing social support to patients on a regular basis to ensure consistency for patients in hospital. Staff discussed offering many kinds of support to patients including financial, mental health, physical activity and signposting to other services and information.

> *‘Providing the support and making sure that it’s regular support and for myself not letting them down, being that reliable… at the end of every session I tell them that I’ll come and see them next week and it’ll be roughly the same time so I’m not going on a ward and they’re not there.’ Participant 12, trained*.

> *‘We’re’ here to try and support you with any issues that you are having, whether that be financial, anything like that we’re sort of signposting services to anything really’. Participant 2, trained*.

### Feedback on MECC conversation starters

Non-trained staff also suggested that before initiating conversations about diet, it would be helpful to first gauge patients’ understanding of healthy eating and weight management. This approach would allow MECC conversations to be better tailored to individual needs, enhancing the relevance of advice provided.

> *‘I suppose in terms of the first question about how important is it for you to eat healthily, I suppose if I was having a conversation I might phrase that slightly different. Because some people might not know what eating healthily is, or eating healthily might be different for different people.’ Participant 7, non-trained*

## Discussion

Quantitative survey findings following MECC training in a mental health setting in the Northeast England suggested that staff perceived improvements in capability and motivation to deliver MECC, with the most notable increase observed in confidence. Confidence increased consistently over time, including among matched participants, suggesting growing self-efficacy with practice. In contrast, perceived social opportunity significantly declined at follow-up (alongside a marginal decline in perceived physical opportunity) in the wider sample, possibly reflecting challenges in sustaining environmental support. These findings were reflected in the qualitative data, where trained staff described greater confidence, enhanced communication skills, and improved ability to tailor conversations to patient needs. Both data sources highlight the role of training in developing psychological capability and reflective motivation, particularly through increased confidence, and strengthened therapeutic relationships. Although reflective motivation slightly declined among matched participants, possibly due to increased awareness of practical limitations. Automatic motivation increased over time, suggesting emerging habits or increased familiarity with delivery. Perceived importance and usefulness of MECC declined significantly at follow-up, which again may indicate that initial enthusiasm was difficult to sustain without continued organisational support. Our qualitative findings show that organisational support and role clarity further facilitated engagement, while persistent barriers, such as time constraints, inconsistent recording systems, and the impact of wider social determinants, were consistently noted across both trained and non-trained groups, mirroring the quantitative decline in physical and social opportunity and reflecting challenges encountered during implementation.

This study highlights how structured MECC training enhances staff capability and confidence to engage in weight-related conversations. Trained staff demonstrated a more systematic, patient-centred approach, employing targeted communication strategies, clear signposting to resources, and increased awareness of when and how to initiate sensitive discussions. In contrast, non-trained staff tended to rely on generalised health promotion strategies and often lacked the confidence or tools to tailor conversations to individual needs. This contrast underscores the value of formal training, particularly in supporting sensitive, context-specific behaviour change communication. The observed differences in staff capability likely reflect training outcomes, linked to the structured and consistent delivery of the MECC programme. A recent fidelity evaluation of MECC in this setting confirmed high adherence to core behaviour change techniques, showing that both the Train the Trainer and Core MECC + AWOYM packages were delivered as intended [21]. Similar training improvements have been reported elsewhere; for example, Bull et al. [22] found that behavioural science interprofessional skills training significantly increased practitioner confidence, competence, and intention to use behaviour change techniques in community health and social care settings.

Despite generally positive perceptions of MECC training, participants made suggestion for both content and broader implementation strategies. Staff (both trained and non-trained) expressed a need for more support in delivering culturally sensitive and individually tailored conversations, particularly when discussing weight-related issues with individuals from diverse backgrounds. These concerns are also echoed by service users themselves. A recent study with service users living with serious mental illness reported limited communication about medication-related weight gain and emphasised the need for more therapeutic, culturally appropriate engagement [23]. Similar gaps were observed in the community: adults taking antidepressants noted that current support often felt generic and disconnected from the psychological and practical challenges they faced [24]. While Kemp et al. [21] confirmed high adherence to core behaviour change techniques in MECC training delivery, wider implementation across the region remains fragmented [5], with delivery often occurring between rather than within organisations, limiting the consistency and routine application of training. Ethnographic research in inpatient mental health settings [25] illustrates how cultural tensions, under-resourcing, and institutional norms further complicate staff’s efforts to promote health behaviours. Consequently, future MECC training may benefit from going beyond generic communication skills to include structured opportunities to practice more sensitive conversations, as suggested by Meade et al. [26] and Nichol, Rodrigues, et al. [6]. It may also be valuable to consider local contextual and cultural adaptations, including approaches already in use such as tailoring MECC delivery to mental health settings [5, 27-28].

Staff consistently highlighted structural and organisational barriers that constrained effective MECC delivery. The use of standardised recording systems could improve continuity of care and follow-up, while socioeconomic disadvantage often made it difficult for service users to act on lifestyle advice, challenges also reflected in secure mental health settings, where fragmented services, obesogenic environments, and poor food provision undermined health promotion efforts [25, 29]. These findings align with feedback from both inpatient [23] and community-based service users [24], who reported that medication side effects, limited cultural awareness, and inconsistent support limited the impact of MECC conversations. Recent evaluations of MECC implementation across Integrated Care Systems (ICS) in England further support this picture, identifying barriers at macro (e.g. funding), organisational (e.g. time, staffing), and individual levels [30]. Meade et al. [15] also found that even when healthcare professionals intend to deliver MECC, environmental constraints, emotional burden, and prioritisation challenges reduce follow-through. Their subsequent qualitative work [26] reinforced the importance of enabling environments, highlighting how factors such as consultation settings, resource visibility, and management support play a pivotal role in embedding MECC into routine care. Together, these findings suggest that beyond enhancing staff capability and motivation, effective MECC implementation depends on coordinated, equity-focused systems that recognise the structural realities of both service users and providers (physical and social opportunity).

### Implications for practice, policy, and future research

Structured MECC training builds staff confidence, communication skills, and ability to tailor weight-related conversations in mental health settings. To enhance this impact, future training could include further behaviour change theory and techniques, as demonstrated by Bull et al. [22], whose behaviour change intervention significantly improved practitioner confidence, competence, and intention to use various behaviour change techniques within conversations with patients. Embedding approaches such as action planning, goal setting, and person-centred conversation skills from the Motivational Interviewing communication approach [31], alongside culturally competent communication, can support staff in initiating sensitive conversations more effectively. Incorporating these elements within MECC aligns with recent calls [32] about the potential of using behavioural science to shape the design, delivery, and evaluation of health promotion training [33].

To strengthen staff capability, MECC training could include experiential learning (e.g. case study analysis), skills rehearsal, and feedback. Embedding behavioural techniques such as action planning (e.g. if-then plans), commitment (e.g. one action they will apply post-training), verbal persuasion, and the use of mnemonics may further enhance reflective and automatic motivation. To support environmental opportunity, implementation must be underpinned by organisational investment, such as protected time, leadership buy-in, and digital infrastructure. Practical strategies could include standardised (but flexible) recording/monitoring MECC delivery systems (e.g. aligned with the 3As elements of Ask, Assist, Act) [5], conversation starters resources, visible reminders (e.g., posters, mouse mats, digital prompts), and peer support channels or messaging groups to foster social support and enable consistent practice across settings.

Future research should extend beyond short-term outcomes and self-reported measures to assess actual changes in practice. This could include observational methods such as follow-up consultations or audio-recordings to examine the use of specific skills and techniques in real-world settings. Larger, longitudinal studies could also help determine the effectiveness of specific teaching methods and whether including particular behaviour change techniques (e.g. action planning) within MECC training enhance the delivery and quality of MECC conversations, and effectively support behaviour change in patients.

### Strengths and limitations

This study offers a detailed exploration of the barriers and facilitators to MECC conversations about weight in a mental health context, drawing on both qualitative and quantitative methods underpinned by the COM-B approach [12]. This research also benefited from collaboration with academic, clinical, and public contributors, supporting the development of contextual relevance. The qualitative component was based on a relatively large and diverse sample, particularly in terms of interprofessional representativeness However, the quantitative survey involved a smaller sample, limiting the generalisability of results and confidence in the observed statistical significance. The sample was self-selected, which may have introduced some bias toward participants with greater interest in MECC, although it included both trained and non-trained staff. Additionally, the study was limited to one NHS trust in Northeast England, which may restrict transferability to other settings. As with similar studies, we assessed short-term outcomes, such as confidence, perceived capability, and motivation, rather than direct measures of behaviour or longer-term practice change. Although planned in our published, pre-registered protocol, the absence of standardised systems to record and monitor MECC delivery limited our ability to assess actual MECC delivery beyond self-report.

## Conclusion

MECC training appeared to improve staff confidence and motivation to engage in health-promoting conversations, though sustained implementation may be challenging due to perceived declines in environmental support and long-term relevance. Findings suggest that refresher training may help sustain perceived opportunity and engagement over time. Together, the findings emphasise the need for sustaining organisational support, role clarity, and reinforcement strategies to embed MECC more fully into routine mental health practice. However, training alone is not a panacea; meaningful implementation must also consider the complex realities of patients’ lives and clinicians’ roles, particularly in relation to mental health and weight-related issues [34].

## Data Availability

The datasets used and/or analysed during the current study are available from the corresponding author on reasonable request.

## Abbreviations

HCP: healthcare professional
MECC: Making Every Contact Count
NHS: National Health Service
SMI: serious mental illness

## Declarations

### Ethics approval and consent to participate

This project was approved by Northumbria University Ethics Approval Committee (ref: 43190). The study was conducted in accordance with the ethical standards of the Declaration of Helsinki and its later amendments. All participants provided consent to enable recording of interviews. Informed consent was obtained from all participants. Consent forms were approved from the above ethics committee. The study and protocol were approved by the R&D department at St Nicholas Hospital, Cumbria, Northumberland, Tyne & Wear NHS Foundation Trust.

### Consent for publication

Not applicable

### Competing Interest

The authors declare that they have no known competing financial interests or personal relationships that could have appeared to influence the work reported in this paper. KM and SF are employed by the healthcare organisation where the study took place and supported this project in terms of recruitment and dissemination.

### Funding

This study/project is funded by the National Institute for Health and Care Research (NIHR) Applied Research Collaboration (ARC) North East and North Cumbria (NIHR200173). The views expressed are those of the author(s) and not necessarily those of the NIHR or the Department of Health and Social Care.

### Author contributions: CRediT

AR, CH, KM, RA, RW, MV, and ML developed the initial study design and secured funding for the study. EK and AR conducted preparation of study materials, data collection and analysis, and drafted summary reports. EK and AR drafted the manuscript and its revised iterations. EK, AR, CH, KM, SF, RA, RW, MV, ML, CR and JH contributed and provided comments on data analysis and interpretation, and report drafts. All co-authors have reviewed and agreed the final draft of the paper submitted for publication. AR acted as guarantor.

## Acknowledgements

We thank all the interviewees who took the time to participate in this research, and the colleagues at CNTW for supporting this project throughout. We would also like to acknowledge Katie Bannister, Penny Spring and Lisa Davies for their support and involvement in the initial funding bid.

## Notes

### Competing Interest Statement

The authors have declared no competing interest.

### Clinical Protocols

https://www.tandfonline.com/doi/full/10.1080/21642850.2023.2174698

