## Supplementary file 1 for "Assessing healthcare professionals’ experiences of delivering opportunistic, weight-related conversations in a mental health setting: a mixed methods study"

MECC Pre training questionnaire

1.1 **1. Please state your age**

________________________________________________________________

1.2 **2. Please state your gender**

- Male (1)
- Female (2)
- Non-binary / third gender (3)
- Prefer not to say (4)

1.3 **3. What is your current role at CNTW? (please select from list)**

▼ Nurse (1) ... Other (13)

Skip To: 1.4 If 1.3 = Other

1.4 **3.1 If you answered 'other'- please state your role**

________________________________________________________________

1.5 **4. How long have you been in this role?**

- 6 months or less (1)
- Over 6 months, up to 1 year (2)
- Over 1 year, up to 3 years (3)
- Over 3 years, up to 5 years (4)
- Over 5 years (5)

1.6 **5. Where is your ward/service located?**

- St. George's Park inpatient (1)
- St. George's Park community (2)
- Northgate inpatient (3)
- Northgate community (4)
- Other (Please state) (5) __________________________________________________

Q34 **6. Please state if you have completed any MECC training prior to this session**

|  | Method of Delivery | | Status | | Training | |
| --- | --- | --- | --- | --- | --- | --- |
|  | Online (1) | In Person (2) | Completed (1) | Not completed (2) | Date of training (Completed) (1) | How often have you completed this training (2) |
| Train the Trainer (1) |  |  |  |  |  |  |
| Core MECC (2) |  |  |  |  |  |  |
| Core MECC + AWOYM (3) |  |  |  |  |  |  |

1.7 **7. Please state what MECC training you are about to receive**

- Train the Trainer (1)
- Core MECC (2)
- Core MECC + AWOYM (A Weight Off Your Mind) (3)

1.8 **8. What is the method of delivery for this training?**

- Online (1)
- In-person (2)

| Page Break |
| --- |

2 **Use of Making Every Contact Count (MECC) + AWOYM** (A Weight Off Your Mind)

 **Please answer the following questions relating to delivering MECC + AWOYM**

2.1 **9. Thinking of the patients you see in a typical week, what percentage of them do you think it is appropriate to deliver MECC + AWOYM to?

 Please drag the button along to answer this question**

|  | 0 | 10 | 20 | 30 | 40 | 50 | 60 | 70 | 80 | 90 | 100 |
| --- | --- | --- | --- | --- | --- | --- | --- | --- | --- | --- | --- |

| Percentage () | 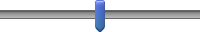 |
| --- | --- |

2.2 **10. Have you ever delivered MECC + AWOYM?**

- No (1)
- Yes (2)

2.3 **11. Of the weekly patients for whom it is appropriate to deliver MECC+ AWOYM, what percentage of them do you usually manage to deliver MECC + AWOYM?** Please drag the button along to answer this question

|  | 0 | 10 | 20 | 30 | 40 | 50 | 60 | 70 | 80 | 90 | 100 |
| --- | --- | --- | --- | --- | --- | --- | --- | --- | --- | --- | --- |

| Percentage () | 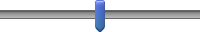 |
| --- | --- |

2.4 **12. Have you delivered MECC+ AWOYM to patients in relation to any of the following behaviours?

 (Please tick all that apply)**

- Smoking (1)
- Drug and alcohol use (2)
- Physical activity (3)
- Healthy eating (4)

2.5 **13. When using MECC + AWOYM, how often do you document the interventions you deliver in a patient's record?**

|  | Never (1) | Sometimes (2) | About half the time (3) | Most of the time (4) | Always (5) |
| --- | --- | --- | --- | --- | --- |
| Please rate (1) |  |  |  |  |  |

2.6 **14. Where do you document MECC + AWOYM that you deliver?

 (Please tick all that apply)**

- MECC Client Record (1)
- Patient paper notes (2)
- Electronic patient records (3)
- RIO (4)
- Other (5)

Skip To: 2.7 If 2.6 = Other

2.7 **15. If you selected 'Other', where do you document MECC + AWOYM?**

________________________________________________________________

2.8 **16. How easy or difficult is it to document MECC + AWOYM?**

|  | Extremely difficult (1) | Somewhat difficult (2) | Neither easy nor difficult (3) | Somewhat easy (4) | Extremely easy (5) |
| --- | --- | --- | --- | --- | --- |
| Please rate (1) |  |  |  |  |  |

| Page Break |
| --- |

3 **Attitudes and motivation towards delivering MECC + AWOYM

Please answer the following questions in relation to the attitudes and motivation you have towards delivering MECC + AWOYM**

3.1 **17. Of the service users you see in a typical working week, with what proportion do you have the PHYSICAL opportunity to Make Every Contact Count?**

 (What is PHYSICAL opportunity? The environment provides the opportunity to engage in the activity concerned. (e.g., sufficient time, the necessary materials, reminders)

|  | 0 | 10 | 20 | 30 | 40 | 50 | 60 | 70 | 80 | 90 | 100 |
| --- | --- | --- | --- | --- | --- | --- | --- | --- | --- | --- | --- |

| Percentage () | 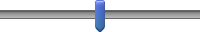 |
| --- | --- |

Q34 **19 (1). Of the service users you see in a typical working week, with what proportion do you have the SOCIAL opportunity to Make Every Contact Count?**

 (What is SOCIAL opportunity?  Interpersonal influences, social cues and cultural norms provide the opportunity to engage in the activity concerned (e.g., other colleagues Making Every Contact Count, support from managers)

|  | 0 | 10 | 20 | 30 | 40 | 50 | 60 | 70 | 80 | 90 | 100 |
| --- | --- | --- | --- | --- | --- | --- | --- | --- | --- | --- | --- |

| Percentage () | 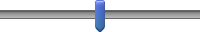 |
| --- | --- |

3.2 **18. I am motivated to Make Every Contact Count**

 (What is motivation? Conscious planning and evaluations (beliefs about what is good and bad) (E.g. I have the desire to, I feel the need to)

|  | Strongly Disagree 1 (1) | 2 (2) | 3 (3) | 4 (4) | 5 (5) | 6 (6) | 7 (7) | 8 (8) | 9 (9) | Strongly Agree 10 (10) |
| --- | --- | --- | --- | --- | --- | --- | --- | --- | --- | --- |
| Please rate (1) |  |  |  |  |  |  |  |  |  |  |

3.3 **19. Making Every Contact Count is something I do automatically**

 (Automatic motivation involves doing something without thinking or having to consciously remember (e.g. ‘is something I do before I realise I’m doing it’)

|  | Strongly Disagree 1 (1) | 2 (2) | 3 (3) | 4 (4) | 5 (5) | 6 (6) | 7 (7) | 8 (8) | 9 (9) | Strongly Agree 10 (10) |
| --- | --- | --- | --- | --- | --- | --- | --- | --- | --- | --- |
| Please rate (1) |  |  |  |  |  |  |  |  |  |  |

3.4 **20. I am PHYSICALLY able to Make Every Contact Count**

 (What is physical capability? Having the physical skill, strength or stamina to engage in the activity concerned. (e.g. I have sufficient physical stamina, I can overcome disability, I have sufficient physical skills)

|  | Strongly Disagree 1 (1) | 2 (2) | 3 (3) | 4 (4) | 5 (5) | 6 (6) | 7 (7) | 8 (8) | 9 (9) | Strongly Agree 10 (10) |
| --- | --- | --- | --- | --- | --- | --- | --- | --- | --- | --- |
| Please rate (1) |  |  |  |  |  |  |  |  |  |  |

3.5 **21. I am PSYCHOLOGICALLY able to Make Every Contact Count**

 (What is psychological capability? Knowledge and/or psychological skills, strength or stamina to engage in the necessary thought processes for the activity concerned. (e.g. having the knowledge, cognitive and interpersonal skills, having the ability to engage in appropriate memory, attention and decision making processes).

|  | Strongly Disagree 1 (1) | 2 (2) | 3 (3) | 4 (4) | 5 (5) | 6 (6) | 7 (7) | 8 (8) | 9 (9) | Strongly Agree 10 (10) |
| --- | --- | --- | --- | --- | --- | --- | --- | --- | --- | --- |
| Please rate (1) |  |  |  |  |  |  |  |  |  |  |

| Page Break |
| --- |

4 **MECC confidence, importance and usefulness

 Please answer the following questions relating to your confidence, perceived importance and usefulness of delivering MECC + AWOYM.**

 Please click one number for each item.

4.1 **22. On a scale of 1-10, how confident do you feel about supporting service users to make behaviour changes? (please click the number)**

|  | Not at all confident 1 (1) | 2 (2) | 3 (3) | 4 (4) | 5 (5) | 6 (6) | 7 (7) | 8 (8) | 9 (9) | Very confident 10 (10) |
| --- | --- | --- | --- | --- | --- | --- | --- | --- | --- | --- |
| Please rate (1) |  |  |  |  |  |  |  |  |  |  |

4.2 **23. On a scale of 1 – 10, how important is it for you to support clients/individuals to make a behaviour change? (please click the number)**

|  | Not at all important 1 (1) | 2 (2) | 3 (3) | 4 (4) | 5 (5) | 6 (6) | 7 (7) | 8 (8) | 9 (9) | Very important 10 (10) |
| --- | --- | --- | --- | --- | --- | --- | --- | --- | --- | --- |
| Please rate (1) |  |  |  |  |  |  |  |  |  |  |

4.3 **24. On a scale of 1 – 10, how useful do you think the conversations you have are at supporting individuals to make a behaviour change? (please click the number)**

|  | Not at all useful 1 (1) | 2 (2) | 3 (3) | 4 (4) | 5 (5) | 6 (6) | 7 (7) | 8 (8) | 9 (9) | Very useful 10 (10) |
| --- | --- | --- | --- | --- | --- | --- | --- | --- | --- | --- |
| Please rate (1) |  |  |  |  |  |  |  |  |  |  |

| Page Break |
| --- |

MECC Post training questionnaire

1 **Some background information about you**

1.1 **1. Please state your age**

________________________________________________________________

1.2 **2. Please state your gender**

▼ Male (1) ... Prefer not to say (4)

1.3 **3. What is your current role at CNTW? (Please select from list)**

▼ Nurse (1) ... Other (13)

Skip To: 1.4 If 3. What is your current role at CNTW? (Please select from list) = Other

1.4 **3.1 If you answered 'Other' please state your role**

________________________________________________________________

1.5 **4. How long have you been in this role?**

- 6 months or less (1)
- Over 6 months, up to 1 year (2)
- Over 1 year, up to 3 years (3)
- Over 3 years, up to 5 years (4)
- Over 5 years (5)

1.6 **5. Where is your ward/ service located? (Please select from list)**

- St. George's Park inpatients (1)
- St. George's Park community (2)
- Northgate inpatients (3)
- Northgate community (4)
- Other (Please state) (5) __________________________________________________

1.7 **6. Please state which MECC training you have received**

- Train the Trainer (1)
- Core MECC (2)
- Core MECC + AWOYM (A Weight Off Your Mind) (3)

1.8 **7. To what extent did you feel satisfied with the training received?**

|  | Extremely dissatisfied (1) | Somewhat dissatisfied (2) | Neither satisfied nor dissatisfied (3) | Somewhat satisfied (4) | Extremely satisfied (5) |
| --- | --- | --- | --- | --- | --- |
| Train the Trainer (1) |  |  |  |  |  |
| Core MECC (2) |  |  |  |  |  |
| Core MECC + AWOYM (3) |  |  |  |  |  |

1.9 **8. What delivery method of training did you receive?**

- Online (1)
- In person (2)

1.10 **9. Overall, how would you rate the method of delivery of training?**

|  | Not effective at all (1) | Slightly effective (2) | Moderately effective (3) | Very effective (4) | Extremely effective (5) |
| --- | --- | --- | --- | --- | --- |
| Online (1) |  |  |  |  |  |
| In-person (2) |  |  |  |  |  |

| Page Break |
| --- |

2 **Use of Making Every Contact Count (MECC) + AWOYM** (A Weight Off Your Mind)

 **Please answer the following questions relating to delivering MECC + AWOYM**

2.1  **10. Thinking of the patients you see in a typical week, what percentage of them do you think it is appropriate to deliver MECC + AWOYM to?**

 Please drag the button along to answer this question

|  | 0 | 10 | 20 | 30 | 40 | 50 | 60 | 70 | 80 | 90 | 100 |
| --- | --- | --- | --- | --- | --- | --- | --- | --- | --- | --- | --- |

| Percentage () | 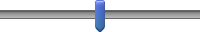 |
| --- | --- |

2.2  **11. Have you ever delivered MECC + AWOYM?**

- Yes (1)
- No (2)

2.3 **12. Of the weekly patients for whom it is appropriate to deliver MECC + AWOYM, what percentage of them do you usually manage to deliver MECC+ AWOYM?**

 Please drag the button along to answer this question

|  | 0 | 10 | 20 | 30 | 40 | 50 | 60 | 70 | 80 | 90 | 100 |
| --- | --- | --- | --- | --- | --- | --- | --- | --- | --- | --- | --- |

| Percentage () | 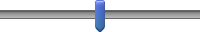 |
| --- | --- |

2.4 **13. Have you delivered MECC + AWOYM to patients in relation to any of the following behaviours?** (Please tick all that apply)

- Smoking (1)
- Drug and Alcohol use (2)
- Physical Activity (3)
- Healthy eating (4)

2.5 **14. When using MECC + AWOYM, how often do you document the interventions you deliver in a patient's record?**

|  | Never (1) | Sometimes (2) | About half the time (3) | Most of the time (4) | Always (5) |
| --- | --- | --- | --- | --- | --- |
| Please rate (1) |  |  |  |  |  |

2.6 **15. Where do you document MECC + AWOYM that you deliver?** (Please tick all that apply)

- MECC Client Record (1)
- Patient paper notes (2)
- Electronic patient records (3)
- RIO (4)
- Other (5)

Skip To: 2.7 If 15. Where do you document MECC + AWOYM that you deliver? (Please tick all that apply) = Other

2.7 **16. If you selected 'other', where do you document MECC + AWOYM?**

________________________________________________________________

2.8 **17. How easy or difficult is it to document MECC + AWOYM?**

|  | Extremely difficult (1) | Somewhat difficult (2) | Neither easy nor difficult (3) | Somewhat easy (4) | Extremely easy (5) |
| --- | --- | --- | --- | --- | --- |
| Please rate (1) |  |  |  |  |  |

2.9 **18. Please describe what makes it easy or difficult for you to document MECC + AWOYM:**

________________________________________________________________

| Page Break |
| --- |

3 **Attitudes and motivation towards delivering MECC + AWOYM

Please answer the following questions in relation to the attitudes and motivation you have towards delivering MECC + AWOYM**

3.1 **19. Of the service users you see in a typical working week, with what proportion do you have the PHYSICAL opportunity to Make Every Contact Count?**

 (What is PHYSICAL opportunity? The environment provides the opportunity to engage in the activity concerned.
 (e.g., sufficient time, the necessary materials, reminders)

|  | 0 | 10 | 20 | 30 | 40 | 50 | 60 | 70 | 80 | 90 | 100 |
| --- | --- | --- | --- | --- | --- | --- | --- | --- | --- | --- | --- |

| Percentage () | 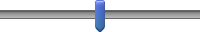 |
| --- | --- |

3.2 **20. Of the service users you see in a typical working week, with what proportion do you have the SOCIAL opportunity to Make Every Contact Count?**

 (What is SOCIAL opportunity? Interpersonal influences, social cues and cultural norms provide the opportunity to engage in the activity concerned (e.g., other colleagues Making Every Contact Count, support from managers)

|  | 0 | 10 | 20 | 30 | 40 | 50 | 60 | 70 | 80 | 90 | 100 |
| --- | --- | --- | --- | --- | --- | --- | --- | --- | --- | --- | --- |

| Percentage () | 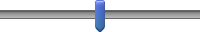 |
| --- | --- |

3.3 **21. I am motivated to Make Every Contact Count**

 (What is motivation? Conscious planning and evaluations (beliefs about what is good and bad) (E.g. I have the desire to, I feel the need to)

|  | Strongly disagree 1 (1) | 2 (2) | 3 (3) | 4 (4) | 5 (5) | 6 (6) | 7 (7) | 8 (8) | 9 (9) | Strongly agree 10 (10) |
| --- | --- | --- | --- | --- | --- | --- | --- | --- | --- | --- |
| Please rate (1) |  |  |  |  |  |  |  |  |  |  |

3.4 **22. Making Every Contact Count is something I do automatically**

 (Automatic motivation involves doing something without thinking or having to consciously remember (e.g. ‘is something I do before I realise I’m doing it’)

|  | Strongly disagree 1 (1) | 2 (2) | 3 (3) | 4 (4) | 5 (5) | 6 (6) | 7 (7) | 8 (8) | 9 (9) | Strongly agree 10 (10) |
| --- | --- | --- | --- | --- | --- | --- | --- | --- | --- | --- |
| Please rate (1) |  |  |  |  |  |  |  |  |  |  |

3.5  **23. I am PHYSICALLY able to Make Every Contact Count**

 (What is physical capability? Having the physical skill, strength or stamina to engage in the activity concerned. (e.g. I have sufficient physical stamina, I can overcome disability, I have sufficient physical skills)

|  | Strongly disagree 1 (1) | 2 (2) | 3 (3) | 4 (4) | 5 (5) | 6 (6) | 7 (7) | 8 (8) | 9 (9) | Strongly agree 10 (10) |
| --- | --- | --- | --- | --- | --- | --- | --- | --- | --- | --- |
| Please rate (1) |  |  |  |  |  |  |  |  |  |  |

3.6 **24. I am PSYCHOLOGICALLY able to Make Every Contact Count**

 **(**What is psychological capability? Knowledge and/or psychological skills, strength or stamina to engage in the necessary thought processes for the activity concerned. (e.g. having the knowledge, cognitive and interpersonal skills, having the ability to engage in appropriate memory, attention and decision making processes).

|  | Strongly disagree 1 (1) | 2 (2) | 3 (3) | 4 (4) | 5 (5) | 6 (6) | 7 (7) | 8 (8) | 9 (9) | Strongly disagree 10 (10) |
| --- | --- | --- | --- | --- | --- | --- | --- | --- | --- | --- |
| Please rate (1) |  |  |  |  |  |  |  |  |  |  |

| Page Break |
| --- |

4. **MECC + AWOYM confidence, importance and usefulness

 Please answer the following questions relating to your confidence, perceived importance and usefulness of delivering MECC + AWOYM.**

 Please click one number for each item.

4.1 **25. On a scale of 1-10, how confident do you feel about supporting service users to make behaviour changes? (please click the number)**

|  | Not at all confident 1 (1) | 2 (2) | 3 (3) | 4 (4) | 5 (5) | 6 (6) | 7 (7) | 8 (8) | 9 (9) | Very confident 10 (10) |
| --- | --- | --- | --- | --- | --- | --- | --- | --- | --- | --- |
| Please rate (1) |  |  |  |  |  |  |  |  |  |  |

4.2 **26. On a scale of 1 – 10, how important is it for you to support clients/individuals to make a behaviour change? (please click the number)**

|  | Not at all important 1 (1) | 2 (2) | 3 (3) | 4 (4) | 5 (5) | 6 (6) | 7 (7) | 8 (8) | 9 (9) | Very important 10 (10) |
| --- | --- | --- | --- | --- | --- | --- | --- | --- | --- | --- |
| Please rate (1) |  |  |  |  |  |  |  |  |  |  |

4.3 **27. On a scale of 1 – 10, how useful do you think the conversations you have are at supporting individuals to make a behaviour change? (please click the number)**

|  | Not at all useful 1 (1) | 2 (2) | 3 (3) | 4 (4) | 5 (5) | 6 (6) | 7 (7) | 8 (8) | 9 (9) | Very useful 10 (10) |
| --- | --- | --- | --- | --- | --- | --- | --- | --- | --- | --- |
| Please rate (1) |  |  |  |  |  |  |  |  |  |  |

| Page Break |
| --- |

MECC Staff Follow up questionnaire

**1. Please state your age**

________________________________________________________________

**2. Please state your gender**

- Male (1)
- Female (2)
- Non-binary / third gender (3)
- Prefer not to say (4)

**3. What is your current role at CNTW? (Please select from list)**

- Nurse (1)
- Nurse associate (2)
- Medic (3)
- AHP- Physio (4)
- AHP- OT (5)
- AHP- SALT (6)
- AHP- Dietician (7)
- Sport/exercise therapist (8)
- Pharmacy (9)
- Students- Discipline (10)
- Healthcare assistant (11)
- Activities coordinator (12)
- Other (Please state) (13) __________________________________________________

**4. How long have you been in this role?**

- 6 months or less (1)
- Over 6 months, up to 1 year (2)
- Over 1 year, up to 3 years (3)
- Over 3 years, up to 5 years (4)
- Over 5 years (5)

**5. Where is your ward/service located?**

- St George's park inpatient (1)
- St George's park community (2)
- Northgate inpatient (3)
- Northgate community (4)
- Other (please state) (5) __________________________________________________

**6. If you have received MECC training, please state which training you have received 

(Please tick all that apply)**

- Train the Trainer (1)
- Core MECC (2)
- Core MECC + AWOYM (A Weight Off Your Mind) (3)
- I have not received MECC training (4)

Skip To: Q33 If 6. If you have received MECC training, please state which training you have received  (Please tic... = I have not received MECC training

**7. To what extent did you feel satisfied with the training received?**

|  | Extremely dissatisfied (1) | Somewhat dissatisfied (2) | Neither satisfied nor dissatisfied (3) | Somewhat satisfied (4) | Extremely satisfied (5) |
| --- | --- | --- | --- | --- | --- |
| Train the Trainer (1) |  |  |  |  |  |
| Core MECC (2) |  |  |  |  |  |
| Core MECC + AWOYM (3) |  |  |  |  |  |

**8. What delivery method of training did you receive?**

- Online (1)
- In person (2)

**9. Overall, how would you rate the method of delivery of training?**

|  | Not effective at all (1) | Slightly effective (2) | Moderately effective (3) | Very effective (4) | Extremely effective (5) |
| --- | --- | --- | --- | --- | --- |
| Online (1) |  |  |  |  |  |
| In person (2) |  |  |  |  |  |

| Page Break |
| --- |

**The next set of questions will ask about MECC in your role.**

 Making Every Contact Count (MECC) Is a public health approach to improving people's health and wellbeing through behaviour change initiated by healthy lifestyle conversations. A Weight Off Your Mind (AWOYM) is a strategy to encourage people with mental health conditions achieve and maintain a healthy weight. 

 The aim of this survey is to evaluate staff who have received MECC+AWOYM training to those who haven't to understand how effective the training is at implementing MECC in the North Locality region.

| Page Break |
| --- |

**Use of Making Every Contact Count (MECC) + AWOYM** (A Weight Off Your Mind)

 **Please answer the following questions relating to delivering MECC + AWOYM**

**10. Thinking of the patients you see in a typical week, what percentage of them do you think it is appropriate to deliver MECC + AWOYM to?**

 Please drag the button along to answer this question

|  | 0 | 10 | 20 | 30 | 40 | 50 | 60 | 70 | 80 | 90 | 100 |
| --- | --- | --- | --- | --- | --- | --- | --- | --- | --- | --- | --- |

| Percentage () | 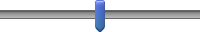 |
| --- | --- |

**11. In the past 8-10 weeks, have you delivered MECC + AWOYM?**

- Yes (1)
- No (2)
- I have NOT received MECC training but HAVE delivered MECC+AWOYM (3)
- I have NOT received MECC training and have NOT delivered MECC + AWOYM (4)

**12. Of the weekly patients for whom it is appropriate to deliver MECC + AWOYM, what percentage of them do you usually manage to deliver MECC+ AWOYM?**

 Please drag the button along to answer this question

|  | 0 | 10 | 20 | 30 | 40 | 50 | 60 | 70 | 80 | 90 | 100 |
| --- | --- | --- | --- | --- | --- | --- | --- | --- | --- | --- | --- |

| Percentage () | 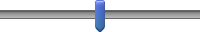 |
| --- | --- |

**13. Have you delivered MECC + AWOYM to patients in relation to any of the following behaviours?** (Please tick all that apply)

- Smoking (1)
- Drug and Alcohol use (2)
- Physical activity (3)
- Healthy eating (4)
- I have not delivered MECC + AWOYM (5)

Skip To: Q19 If 13. Have you delivered MECC + AWOYM to patients in relation to any of the following behaviours? (... = I have not delivered MECC + AWOYM

**14. When using MECC + AWOYM, how often do you document the interventions you deliver in a patient's record?**

|  | Never (1) | Sometimes (2) | About half the time (3) | Most of the time (4) | Always (5) |
| --- | --- | --- | --- | --- | --- |
| Click to write Statement 1 (1) |  |  |  |  |  |

**15. Where do you document MECC + AWOYM that you deliver?** (Please tick all that apply)

- MECC Client record (1)
- Patient paper notes (2)
- Electronic patient records (3)
- RIO (4)
- Other (Please state) (5) __________________________________________________

**16. How easy or difficult is it to document MECC + AWOYM?**

|  | Extremely difficult (1) | Somewhat difficult (2) | Neither easy nor difficult (3) | Somewhat easy (4) | Extremely easy (5) |
| --- | --- | --- | --- | --- | --- |
| Please rate (1) |  |  |  |  |  |

**17. Please describe what makes it easy or difficult for you to document MECC + AWOYM:**

________________________________________________________________

| Page Break |
| --- |

**Attitudes and motivation towards delivering MECC + AWOYM

 Please answer the following questions in relation to the attitudes and motivation you have towards delivering MECC + AWOYM**

**18. Of the service users you see in a typical working week, with what proportion do you have the PHYSICAL opportunity to Make Every Contact Count?**

 (What is PHYSICAL opportunity? The environment provides the opportunity to engage in the activity concerned. (e.g., sufficient time, the necessary materials, reminders)

|  | 0 | 10 | 20 | 30 | 40 | 50 | 60 | 70 | 80 | 90 | 100 |
| --- | --- | --- | --- | --- | --- | --- | --- | --- | --- | --- | --- |

| Percentage () | 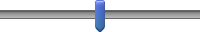 |
| --- | --- |

**19. Of the service users you see in a typical working week, with what proportion do you have the SOCIAL opportunity to Make Every Contact Count?**

 (What is SOCIAL opportunity? Interpersonal influences, social cues and cultural norms provide the opportunity to engage in the activity concerned (e.g., other colleagues Making Every Contact Count, support from managers)

|  | 0 | 10 | 20 | 30 | 40 | 50 | 60 | 70 | 80 | 90 | 100 |
| --- | --- | --- | --- | --- | --- | --- | --- | --- | --- | --- | --- |

| Percentage () | 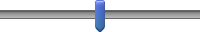 |
| --- | --- |

**20. I am motivated to Make Every Contact Count**

 (What is motivation? Conscious planning and evaluations (beliefs about what is good and bad) (E.g. I have the desire to, I feel the need to)

|  | Strongly disagree 1 (1) | 2 (2) | 3 (3) | 4 (4) | 5 (5) | 6 (6) | 7 (7) | 8 (8) | 9 (9) | Strongly agree 10 (10) |
| --- | --- | --- | --- | --- | --- | --- | --- | --- | --- | --- |
| Pleas rate (1) |  |  |  |  |  |  |  |  |  |  |

**21. Making Every Contact Count is something I do automatically**

 Automatic motivation involves doing something without thinking or having to consciously remember (e.g. ‘is something I do before I realise I’m doing it’)

|  | Strongly disagree 1 (1) | 2 (2) | 3 (3) | 4 (4) | 5 (5) | 6 (6) | 7 (7) | 8 (8) | 9 (9) | Strongly agree 10 (10) |
| --- | --- | --- | --- | --- | --- | --- | --- | --- | --- | --- |
| Please rate (1) |  |  |  |  |  |  |  |  |  |  |

**22. I am PHYSICALLY able to Make Every Contact Count**

 (What is physical capability? Having the physical skill, strength or stamina to engage in the activity concerned. (e.g. I have sufficient physical stamina, I can overcome disability, I have sufficient physical skills)

|  | Strongly disagree 1 (1) | 2 (2) | 3 (3) | 4 (4) | 5 (5) | 6 (6) | 7 (7) | 8 (8) | 9 (9) | Strongly agree 10 (10) |
| --- | --- | --- | --- | --- | --- | --- | --- | --- | --- | --- |
| Please rate (1) |  |  |  |  |  |  |  |  |  |  |

**23. I am PSYCHOLOGICALLY able to Make Every Contact Count**

 (What is psychological capability? Knowledge and/or psychological skills, strength or stamina to engage in the necessary thought processes for the activity concerned. (e.g. having the knowledge, cognitive and interpersonal skills, having the ability to engage in appropriate memory, attention and decision making processes).

|  | Strongly disagree 1 (1) | 2 (2) | 3 (3) | 4 (4) | 5 (5) | 6 (6) | 7 (7) | 8 (8) | 9 (9) | Strongly agree 10 (10) |
| --- | --- | --- | --- | --- | --- | --- | --- | --- | --- | --- |
| Please rate (1) |  |  |  |  |  |  |  |  |  |  |

| Page Break |
| --- |

**MECC + AWOYM confidence, importance and usefulness**

 Please answer the following questions relating to your confidence, perceived importance and usefulness of delivering MECC + AWOYM.

 Please click one number for each item.

**24. On a scale of 1-10, how confident do you feel about supporting service users to make behaviour changes?** (please click the number)

|  | Not at all confident 1 (1) | 2 (2) | 3 (3) | 4 (4) | 5 (5) | 6 (6) | 7 (7) | 8 (8) | 9 (9) | Very confident 10 (10) |
| --- | --- | --- | --- | --- | --- | --- | --- | --- | --- | --- |
| Click to write Statement 1 (1) |  |  |  |  |  |  |  |  |  |  |

**26. On a scale of 1 – 10, how important is it for you to support clients/individuals to make a behaviour change?** (please click the number)

|  | Not at all important 1 (1) | 2 (2) | 3 (3) | 4 (4) | 5 (5) | 6 (6) | 7 (7) | 8 (8) | 9 (9) | Very important 10 (10) |
| --- | --- | --- | --- | --- | --- | --- | --- | --- | --- | --- |
| Click to write Statement 1 (1) |  |  |  |  |  |  |  |  |  |  |

**26. On a scale of 1 – 10, how useful do you think the conversations you have are at supporting individuals to make a behaviour change?** (please click the number)

|  | Not at all useful (1) | Slightly useful (2) | Moderately useful (3) | Very useful (4) | Extremely useful (5) |
| --- | --- | --- | --- | --- | --- |
| Click to write Statement 1 (1) |  |  |  |  |  |

| Page Break |
| --- |

**27. Please reflect on the following statements about using MECC in your role.** Circle one response between 1 (completely disagree) to 5 (completely agree).

|  | Strongly disagree (1) | Somewhat disagree (2) | Neither agree nor disagree (3) | Somewhat agree (4) | Strongly agree (5) |
| --- | --- | --- | --- | --- | --- |
| Using MECC meets my approval (1) |  |  |  |  |  |
| Using MECC is appealing to me (2) |  |  |  |  |  |
| Using MECC seems fitting (3) |  |  |  |  |  |
| Using MECC seems suitable (4) |  |  |  |  |  |
| Using MECC seems applicable (5) |  |  |  |  |  |
| Using MECC seems like a good match (6) |  |  |  |  |  |
| Using MECC seems implementable (7) |  |  |  |  |  |
| Using MECC seems possible (8) |  |  |  |  |  |
| Using MECC seems doable (9) |  |  |  |  |  |
| Using MECC seems easy to use (10) |  |  |  |  |  |
| I like MECC (11) |  |  |  |  |  |
| I welcome MECC (12) |  |  |  |  |  |
