## Supplementary file 2 for "Assessing healthcare professionals’ experiences of delivering opportunistic, weight-related conversations in a mental health setting: a mixed methods study"

**MECC approach at CNTW Interview topic guide (trained staff)**

Thank you for agreeing to take part in this interview. We are inviting a number of health care professionals involved in delivering the MECC approach at CNTW so I am pleased you are able to come along and help. We are interested in your behaviour (in delivering the MECC approach), your perception of the impact on patients and ways to help more people in the future.

1. **DELIVERING THE MECC APPROACH:**

**Intentions and goals**

Why are you interested in delivering the MECC approach, what motivates you (relatives, and friends, community, making a difference)?

What do you hope to achieve by delivering the MECC approach?

**Reinforcement**

What feedback do you receive from the MECC approach? How rewarding is delivering this approach?

Do you receive any feedback from patients after delivering the MECC approach? If so what format is this in? What are their general thoughts?

How could the MECC approach be improved?

**Knowledge**

What do you need to know to deliver the MECC approach? Do you feel you need to know more about the MECC approach?

What training did you receive? Would you be interested in receiving MECC training? (why/why not)

Was the training you received enough? What has prevented you from receiving MECC training? Or has there been opportunity to take part in MECC training?

Do you need refresher training?

Did the training include watching other providers? If so what did you learn?

How is it promoted in your hospital? Has MECC training been promoted in your hospital/work setting?

**Memory. attention and decision processes**

Is delivering a brief lifestyle advice something you were doing already? Is this a new behaviour? If existing, how do you make it a habit? How can it be facilitated?

**Social influences on you as a trainer**

Thinking about your organisation, what factors might enable (or inhibit) the delivery of the MECC approach? (social, cultural, time, gender)

**Beliefs about capabilities and skills**

How confident are you in your skill and capability to deliver the MECC approach?

Are you more confident in some areas (activities) than others?

What are the challenges in your work?

**Optimism**

How confident are you that the problems of implementing the MECC approach will be solved in the future?

**Behavioural regulation, attention and decision processes**

Are there procedures or ways of working that encourage the delivery of the MECC approach?

How do you decide what to do in each session? Do you remember everything you should / could show people?

How do you prepare for each session?

**Environmental context and resources**

Are you supported to implement the MECC approach? Does this allow you to do so as intended?

Are there competing tasks and time constraints? If so what are these?

Are the necessary resources available to those expected to deliver the MECC approach? What are the necessary resources?

**Social role and identity**

What do you do as a deliverer of the MECC approach?

Is delivering the MECC approach compatible or in conflict with professional standards/identity?

**Emotion**

To what extent do emotional factors facilitate or hinder the delivery of the MECC approach?

**MECC APPROACH IMPACT: your perceptions of your service users**

**Intentions and goals**

What do you think influences patients to engage with the MECC approach?

What do you think are the important barriers and facilitators for patients engaging with the MECC approach?

**Reinforcement**

What do you think helps patients to stay motivated?

**Social influences on your clients**

How is your delivery affected by social and cultural factors of patients you wish to engage with the MECC approach?

**Beliefs about capabilities and skills**

Do you think the information provided in the MECC approach is adequate?

Closing question: Is there anything you would like to mention that you don’t think we have covered?

**MECC approach at CNTW Interview topic guide (Non-trained staff)**

Thank you for agreeing to take part in this interview. We are inviting a number of health care professionals involved in delivering the MECC approach at CNTW so I am pleased you are able to come along and help. We are interested in your behaviour (in delivering the MECC approach), your perception of the impact on patients and ways to help more people in the future.

2. **DELIVERING THE MECC APPROACH:**

**Intentions and goals**

What do you know about MECC? Show example of MECC script- include screening question on sign up form (do you have 1-1 contact with patients) send email prior to interview.

MECC is-

Tell me about your role and the patients you see?

Do you have 1-1 contact with patients?

How do you plan sessions?

Is providing behaviour change/healthy lifestyle advice something you do as part of your role?

Are you interested in knowing more about the MECC approach?

Is the MECC approach used by other staff in your department?

Training- have you heard about MECC training?

What MECC training has been advertised to you?

Would you like to take part in MECC training?

What barriers have prohibited you from taking part in MECC training?

Have you taken part in any other training?

Was this more applicable to your role?

Would you like to know more about MECC?

Would you be supported/encouraged to take part in MECC training if you wanted to?

Would you be supported to use the MECC approach?

What are the main challenges in your role?

How engaged are patients with you?

Do you think they would benefit from brief lifestyle advice?

What are your views on having MECC conversations with patients?

Any other questions to add?

Why are you interested in delivering the MECC approach, what motivates you (relatives, and friends, community, making a difference)?

What do you hope to achieve by delivering the MECC approach?

**Reinforcement**

What feedback do you receive from the MECC approach? How rewarding is delivering this approach?

Do you receive any feedback from patients after delivering the MECC approach? If so what format is this in? What are their general thoughts?

How could the MECC approach be improved?

**Knowledge**

What do you need to know to deliver the MECC approach? Do you feel you need to know more about the MECC approach?

What training did you receive? Would you be interested in receiving MECC training? (why/why not)

Was the training you received enough? What has prevented you from receiving MECC training? Or has there been opportunity to take part in MECC training?

Do you need refresher training?

Did the training include watching other providers? If so what did you learn?

How is it promoted in your hospital? Has MECC training been promoted in your hospital/work setting?

**Memory. attention and decision processes**

Is delivering a brief lifestyle advice something you were doing already? Is this a new behaviour? If existing, how do you make it a habit? How can it be facilitated?

**Social influences on you as a trainer**

Thinking about your organisation, what factors might enable (or inhibit) the delivery of the MECC approach? (social, cultural, time, gender)

**Beliefs about capabilities and skills**

How confident are you in your skill and capability to deliver the MECC approach?

Are you more confident in some areas (activities) than others?

What are the challenges in your work?

**Optimism**

How confident are you that the problems of implementing the MECC approach will be solved in the future?

**Behavioural regulation, attention and decision processes**

Are there procedures or ways of working that encourage the delivery of the MECC approach?

How do you decide what to do in each session? Do you remember everything you should / could show people?

How do you prepare for each session?

**Environmental context and resources**

Are you supported to implement the MECC approach? Does this allow you to do so as intended?

Are there competing tasks and time constraints? If so what are these?

Are the necessary resources available to those expected to deliver the MECC approach? What are the necessary resources?

**Social role and identity**

What do you do as a deliverer of the MECC approach?

Is delivering the MECC approach compatible or in conflict with professional standards/identity?

**Emotion**

To what extent do emotional factors facilitate or hinder the delivery of the MECC approach?

**MECC APPROACH IMPACT: your perceptions of your service users**

**Intentions and goals**

What do you think influences patients to engage with the MECC approach?

What do you think are the important barriers and facilitators for patients engaging with the MECC approach?

**Reinforcement**

What do you think helps patients to stay motivated?

**Social influences on your clients**

How is your delivery affected by social and cultural factors of patients you wish to engage with the MECC approach?

**Beliefs about capabilities and skills**

Do you think the information provided in the MECC approach is adequate?

Closing question: Is there anything you would like to mention that you don’t think we have covered?
